# A composite socioeconomic deprivation index from *All of Us* survey data: associations with health outcomes and disparities

**DOI:** 10.1101/2024.10.04.24314904

**Authors:** Sonali Gupta, Vincent Lam, I. King Jordan, Leonardo Mariño-Ramírez

## Abstract

Socioeconomic deprivation – defined as a lack of social, economic and material resources – is associated with poor health outcomes and health disparities between population groups. The *All of Us* Research Program is a longitudinal cohort study of diverse participants from the United States, with demographic and social determinants of health data gleaned from participant surveys and health outcome data derived from electronic health records. We developed a composite index of socioeconomic deprivation (iSDI) using a cohort of 202,919 *All of Us* participants – based on education, employment, health insurance, housing, and income data – and we associated iSDI with health outcomes and disparities. iSDI is significantly associated with 970 out of 1,755 (55.3%) health conditions modeled here, with 661 positive and 309 negative associations. Mental disorders and circulatory diseases show the highest proportion of positive associations with iSDI, whereas neoplasms and congenital anomalies show the highest proportion of negative associations. Black (0.55) and Hispanic (0.52) *All of Us* participants show higher average iSDI values compared to White (0.29) and Asian (0.24) participants; although the majority of iSDI variation is found within (76.8%) rather than between (23.2%) groups. iSDI mediates 213 out of 399 (53.5%) Black health disparity conditions and 173 out of 297 (58.2%) Hispanic health disparity conditions. The composite socioeconomic deprivation index (iSDI) developed here is associated with a wide variety of health outcomes and disparities in the *All of Us* cohort, and we make participant iSDI values available on the Researcher Workbench to support future studies on social determinants of health.

## INTRODUCTION

Socioeconomic deprivation refers to a lack of the social, economic, and material resources that are required to ensure overall well-being and standard of living [1]. Socioeconomic deprivation can be measured by a variety of factors, including education, employment, income, and housing [2–5]. Numerous health outcomes, and in particular health disparities between social groups, have been associated with socioeconomic deprivation [6–11]. Socioeconomic deprivation can affect health outcomes via limited access to healthcare, elevated stress, unhealthy behaviors, and harmful environmental exposures.

The All of Us Research Program (*All of Us* hereafter) is a population biobank of United States residents that includes participant health outcome data gleaned from electronic health records along with demographic and social determinants of health data taken from participant surveys [12–14]. The current *All of Us* cohort includes data from more than 400,000 participants, with outcomes for thousands of disease diagnoses, and the program aims to recruit one million participants. Together, *All of Us* participant health record and survey data provide an opportunity to evaluate associations of socioeconomic deprivation with health outcomes and disparities for numerous diseases.

Socioeconomic deprivation is often measured at the area-level using composite indices that combine measures across multiple dimensions of deprivation [15]. The *All of Us* Researcher Workbench provides a composite metric of area-based socioeconomic deprivation, with values assigned based on participants’ zip codes (zSDI). The zSDI values are taken from the nationwide community deprivation index, which combines socioeconomic variables on education, health insurance, housing, income, and poverty from the American Community Survey [5,16]. *All of Us* currently assigns participants three-digit zip codes that cover relatively large geographic areas compared to more granular five-digit zip codes or census tracts. This may result in participants with different individual-levels of socioeconomic deprivation being assigned the same area-based zSDI values, particularly for diverse urban and suburban communities, thereby potentially limiting the utility of zSDI for modelling health outcomes and disparities in the *All of Us* cohort. The derivation of an analogous individual-level socioeconomic deprivation index for *All of Us* participants could help to overcome this limitation.

The objectives of this study were (i) to derive a quantitative measure of individual-level socioeconomic deprivation using *All of Us* survey questions and (ii) to analyze associations of the derived socioeconomic deprivation metric with health outcomes and disparities in the *All of Us* cohort. We used *All of Us* participant survey data to calculate an individual-level socioeconomic deprivation index (iSDI), analogous to the currently available area-based zSDI, which combines information on participant education, employment, health insurance, housing, and income. We then conducted a series phenome-wide screens of the *All of Us* cohort to evaluate associations of iSDI with health outcomes, to compare the effects of individual-level iSDI versus area-based zSDI on health outcomes, and to evaluate the extent to which iSDI explains observed racial and ethnic health disparities

## MATERIALS AND METHODS

### Individual-level socioeconomic deprivation index (iSDI)

We derived an individual-level socioeconomic deprivation index (iSDI) for *All of Us* participants based on answers to questions from the “Basics Survey” that participants fill out upon enrollment. The following survey questions were used, each of which covers a specific socioeconomic variable: (1) Education – “What is the highest level of education you have completed or the last year of school you attended?”, (2) Employment – “What is your current employment situation?”, (3) Health Insurance – “Do you have health insurance or another form of healthcare coverage?”, (4) Housing – “Do you own or rent your place of residence?”, and (5) Income – “What is your total annual household income from all sources?”. The questions and responses for education, employment, housing, and income are based on the Behavioral Risk Factor Surveillance System (BRFSS), an annual national health-related telephone survey conducted by the Centers for Disease Control and Prevention (CDD) [17]. The question and responses for health insurance are based on the National Health and Nutrition Examination Survey (NHANES), a program conducted by the National Center for Health Statistics that aims to assess the health and nutritional status of the Unites States population [18].

Participant responses to each of these questions were coded as ordinal values, where higher ordinal values correspond to greater socioeconomic deprivation (Supplementary Table 1 and 2). For questions where the order of responses was ambiguous, total annual income was used as the primary indicator of deprivation and the order for other features was refined by maximizing the spearman correlation of each to the order of the income responses. Multivariate Imputation by Chained Equations (MICE) from the *R* package “*mice*” was used to impute missing question-response data [19]. The proportional odds model coded by “*polr*” (ordered, >2 levels) was used to impute all features except insurance for which logreg, logistic regression imputation (binary data, factor with 2 levels) was used. The number of desired imputed datasets (“*m*”) was kept at 5 and the maximum number of iterations (“maxint”) was set to 20. Validation was performed by leaving out 9-10% of observed participant question-response values, imputing these pseudo-missing values, and then checking the correspondence between the observed values and the imputed pseudo-missing values. The imputed dataset that showed the highest percent matching with known data was taken as the final participant question-response ordinal dataset.

Principal component analysis (PCA) of the final participant question-response ordinal dataset was used to calculate the composite iSDI. Polychoric correlations were calculated based on participant responses to all pairs of questions using the function “*polychoric*” from the *R* package “*psych*” [20]. PCA eigenvalues and eigenvectors were calculated from the resulting polychoric correlation matrix using the R function “*eigen*”. We selected the first principal component (PC) to calculate the iSDI based on the percentage of variance explained across PCs. Participant values from PC1 were scaled from 0 to 1 such that a higher value corresponds to higher deprivation level. The iSDI value for participant i is calculated as:

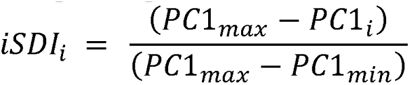

where PC1_max_ and PC1_min_ are the maximum and minimum values of PC1 for all participants.

### *All of Us* study cohort

Our study cohort is made of up volunteer participants from the NIH *All of Us* Research Program (*All of Us*). Participants enrolled in the program electronically or through a participating healthcare provider. *All of Us* inclusion criteria included adults aged 18 and above living in the United States or its territories. Exclusion criteria included individuals who were incarcerated or unable to provide informed consent. Participants provided informed consent for research, and the *All of Us* operational protocol was approved by the NIH Institutional Review Board (#2016-05). *All of Us* participant data were accessed from version 7 of the registered tier dataset, and all analyses were performed using the *All of Us* Researcher Workbench [21].

Participant survey, demographic, and health outcome data were used for this study. Participant responses from the “Basics Survey” were analyzed as described in the previous section. Participant demographic data on age, sex, race, and ethnicity were also taken from the “Basics Survey”. For race and ethnicity, participants were asked “Which categories describe you?”: (1) American Indian or Alaska Native, (2) Asian, (3) Black, African American, or African, (4) Hispanic, Latino, or Spanish, (5) Middle Eastern or North African, (6) Native Hawaiian or other Pacific Islander, (7) White. *All of Us* considers Hispanic, Latino, or Spanish as an ethnic group and all other groups as racial groups, based on the current Office of Management and Budget (OMB) Standards for the Classification of Federal Data on Race and Ethnicity. Information regarding participants who chose American Indian or Alaska Native is currently not available on the *All of Us* Researcher Workbench. Our study cohort is focused individuals from the four most numerous racial and ethnic groups in the *All of Us* cohort: Asian, Black, African American, or African (Black hereafter), Hispanic, Latino, or Spanish (Hispanic hereafter), and White.

Data on *All of Us* participant health outcomes were taken from participants’ electronic health record data recorded by International Classification of Diseases Version 10 (ICD-10) and International Classification of Diseases Version 9 (ICD-9) codes [22]. ICD codes were converted to phecodes, which scale well to biobank-size datasets and are widely used to define disease phenotypes from electronic health record data, for health outcome modeling [23]. Phecodes serve to group granular ICD codes into unified diagnosis codes to define cases and specify exclusion criteria for closely related conditions to define controls.

### Health outcome and disparity modeling

Statistical analyses were performed using the *stats* package, and regression analyses were performed using the *glm* function, with R version 4.2.2. Statistical plots were created using the *ggplot2* R package [24]. Health outcome and disparity modeling were performed via multivariable logistic regression. Health outcomes (dependent variables) were modeled as cases (1) or controls (0) based on phecode data, and model predictors (independent variables) included socioeconomic deprivation, race, and ethnicity. Age and sex were included as covariates in all models. Attenuation of race and ethnicity effects on health outcomes by socioeconomic deprivation was quantified by comparing race and ethnicity effect size estimates for unadjusted models (β_unadjusted_) and for models adjusted by socioeconomic deprivation (β_adjusted_). The proportion of the race and ethnicity effect sizes attenuated by socioeconomic deprivation was calculated as: (β_unadjusted_ – β_adjusted_) β_unadjusted_. The significance of the differences between unadjusted and adjusted effects sizes were measured using the Students t-test, with Bonferroni correction for multiple tests. Mediation analysis was performed using the mediation R package version 4.5.0 [25].

## RESULTS

### Individual-level socioeconomic deprivation index (iSDI)

We derived an individual-level socioeconomic deprivation index (iSDI) from *All of Us* participant survey data using the approach detailed in the Materials and Methods section. The iSDI is a composite metric of participant socioeconomic deprivation, which includes information from survey questions on education, employment, health insurance, housing, and income. Participant responses to these five questions were encoded as ordinal values, with higher values corresponding to greater socioeconomic deprivation (Supplementary Table 1). These questions capture distinct but related dimensions of participant socioeconomic deprivation, with answers to all pairs of questions positively correlated across participants. For some questions, employment and health insurance in particular, there was ambiguity in terms of how responses could be ordered from lowest to highest socioeconomic deprivation. Six different ordering schemes were evaluated to assess how robust the correlations were to different possible response orderings and to choose the optimal ordering scheme (Supplementary Table 2). Changes in response ordering did not have a large effect on the median of the pairwise Spearman correlations, and we chose the ordering with the highest median correlation as the optimal scheme for subsequent analysis (Supplementary Figure 1). For the optimal ordering scheme, Spearman correlations range from ρ=0.17 for employment and health insurance to ρ=0.61 for education and income (Figure 1A and Supplementary Figure 2). Income shows the highest correlation values across all variables. Pairwise correlations were calculated with (Figure 1A) and without (Supplementary Figure 2) imputation of missing response data. Imputation improves the pairwise correlations slightly, and the imputed data were used for subsequent analysis.

**Figure 1.**
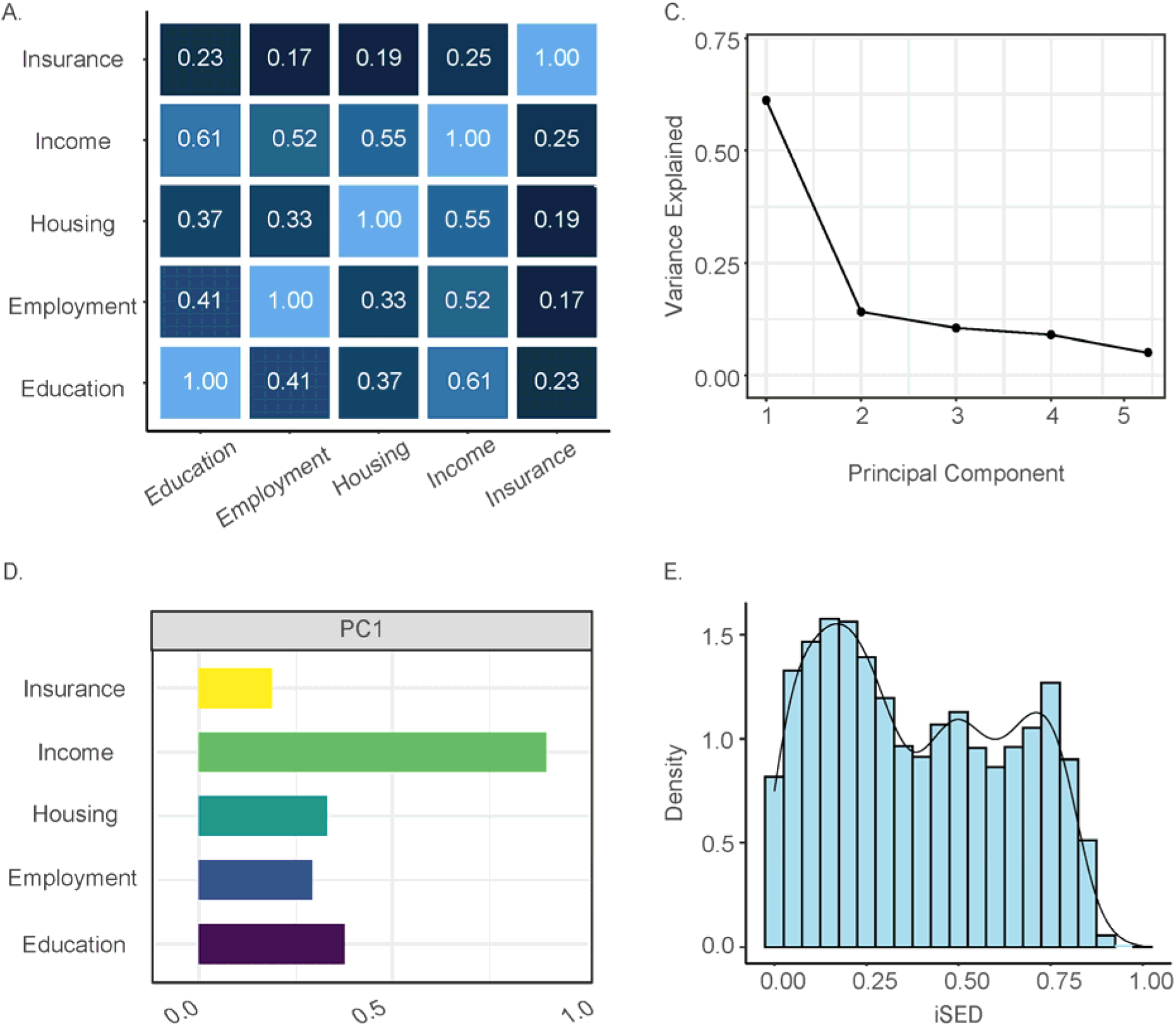
Individual-level socioeconomic deprivation index (iSDI). (A) Pairwise Spearman rank (ρ) correlations between the five constituent socioeconomic deprivation measures taken from the *All of Us* participant survey. (B) Percent variance explained by each principal component (PC). (C) PC1 loadings for each socioeconomic deprivation measure. (D) Distribution of the normalized iSDI metric based on PC1 values.

These correlations indicate that using any one of these variables alone will not capture all aspects of participant socioeconomic deprivation, while using two or more of the variables could lead to multicollinearity in health outcome association models. To facilitate modelling health outcomes by socioeconomic deprivation, principal component analysis (PCA) was used to reduce the dimensionality of the participant survey response data and to generate a composite socioeconomic deprivation index that explains most of variation in the data. The first principal component (PC) explains 61.18% of the variance in the participant survey data, followed by 14.13% and 10.54% for PC2 and PC3, respectively (Figure 1B). The variable loadings for PC1 are all positive, with greater values indicating higher socioeconomic deprivation (Figure 1C). Income shows the greatest effect on socioeconomic deprivation as measured by PC1. Given the large amount of variance explained, and the consistent variable loading values, PC1 values were chosen for the *All of Us* participant iSDI. PC1 values were max-min normalized to yield a distribution of iSDI values from 0 to 1, with higher values representing greater socioeconomic deprivation (Figure 1D).

We compared our individual-level iSDI to the area-based community deprivation index (zSDI), which is currently provided for *All of Us* participants. The zSDI is also a composite metric based on education, health insurance, housing, income, and poverty [5], but values of this index are assigned to *All of Us* participants based on their zip codes, with all participants living in the same zipcode assigned the same value. This may result in participants with different levels of socioeconomic deprivation being assigned the same values, particularly for diverse urban and suburban communities, which is likely exacerbated by the fact that *All of Us* currently uses high-level, three-digit zip codes to participants. Participant values of the individual-level iSDI derived here are positively correlated with values of the area-based zSDI currently used by *All of Us* (Pearson correlation r=0.28).

### iSDI and health outcomes

We associated the iSDI metric derived here with 1,755 diseases (conditions), controlling for age and sex, to evaluate the relationship between socioeconomic deprivation and health outcomes for a cohort of 202,923 *All of Us* participants (Table 1). iSDI was found to be significantly associated with 970 out of 1,755 (55.3%) diseases after controlling for multiple tests (Bonferroni adjusted p<2.85×10^−5^; Figure 2). iSDI is positively associated with 661 (68.1%) and negatively associated with 309 (31.9%) diseases (Table 2). Mental disorders (96.3% positive) and circulatory diseases (90.9% positive) have the highest proportion of positive iSDI associations, whereas neoplasms (88.2% negative) and congenital anomalies (87.5% negative) have the highest proportion of negative iSDI associations.

**Figure 2.**
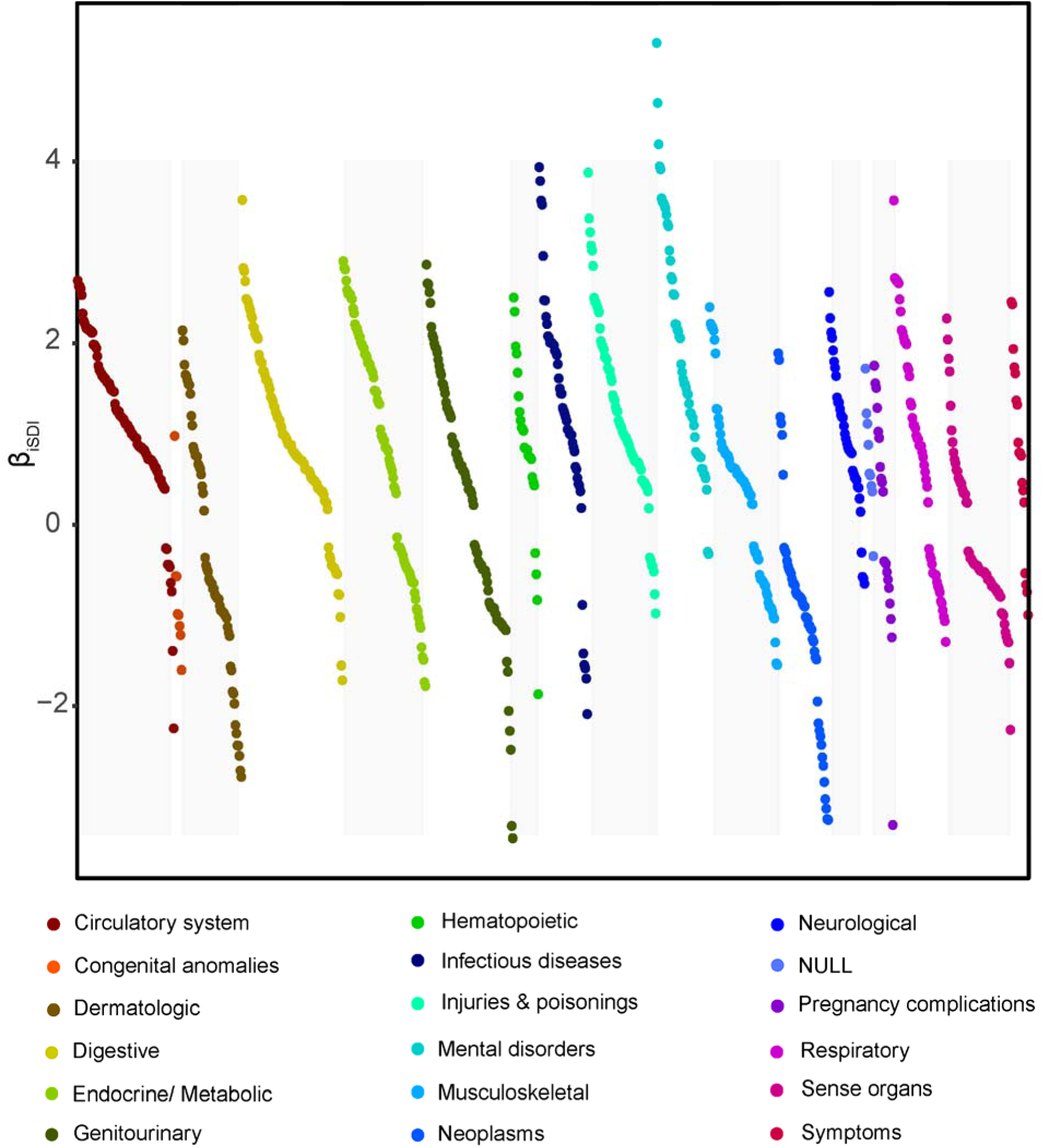
iSDI-disease associations. 1,755 diseases (conditions) were modeled with multivariable logistic regression as: Disease ∼ iSDI + Age + Sex. The effect sizes (β_iSDI_) for all statistically significant associations (Bonferroni adjusted p<2.85×10^−5^) are shown on the y-axis, with related diseases grouped into phecode categories as indicated.

**Table 1.** Study cohort characteristics.

|  | **Full Cohort** | **Asian** | **Black** | **Hispanic** | **White** |
| --- | --- | --- | --- | --- | --- |
| n (%) | 202,919<br>(100%) | 5,373<br>(2.65%) | 40,259<br>(19.84%) | 40,762<br>(20.09%) | 116,525<br>(57.42%) |
| Male n (%) | 75,522<br>(37.22%) | 2,011<br>(37.43%) | 15,504<br>(38.51%) | 12,507<br>(30.68%) | 45,500<br>(39.05%) |
| Female n (%) | 127,397<br>(62.78%) | 3,362<br>(62.57%) | 24,755<br>(61.49%) | 28,255<br>(69.32%) | 71,025<br>(60.95%) |
| iSDI mean (sd) | 0.39 (0.25) | 0.24 (0.19) | 0.55 (0.22) | 0.52 (0.22) | 0.29 (0.22) |

**Table 2.** iSDI-disease associations. Numbers of significantly positive and negative iSDI-disease associations are shown for each phecode disease (condition) category. The percentages of positive and negative associations are shown for each category. Model specification: Disease ∼ iSDI + age + sex.

| Disease (condition) category | iSDI-disease associations |  |
| --- | --- | --- |
|  | Positive | Negative |
| Circulatory | 90 (90.9%) | 9 (9.1%) |
| Congenital anomalies | 1 (12.5%) | 7 (87.5%) |
| Dermatologic | 23 (37.7%) | 38 (62.3%) |
| Digestive | 88 (85.4%) | 15 (14.6%) |
| Endocrine / Metabolic | 55 (64.7%) | 30 (35.3%) |
| Genitourinary | 49 (55.1%) | 40 (44.9%) |
| Hematopoietic | 22 (84.6%) | 4 (15.4%) |
| Infectious diseases | 44 (88.0%) | 6 (12.0%) |
| Injuries and poisonings | 63 (90.0%) | 7 (10.0%) |
| Mental disorders | 52 (96.3%) | 2 (3.7%) |
| Musculoskeletal | 45 (64.3%) | 25 (35.7%) |
| Neoplasms | 6 (11.8%) | 45 (88.2%) |
| Neurological | 33 (89.2%) | 4 (10.8%) |
| Null | 8 (88.9%) | 1 (11.1%) |
| Pregnancy complications | 10 (50.0%) | 10 (50.0%) |
| Respiratory | 36 (66.7%) | 18 (33.3%) |
| Sense Organs | 22 (33.3%) | 44 (66.7%) |
| Symptoms | 14 (77.8%) | 4 (22.2%) |
| Total | 661 (68.1%) | 309 (31.9%) |

Schizophrenia (β=5.31, p≈0), mental retardation (β=4.64, p=5.36×10^−87^), and substance addiction (β=4.19, p≈0) are the diseases with the highest positive iSDI associations. Azoospermia and oligospermia (β=-3.46, p=5.72×10^−17^), male infertility (β=-3.32, p=6.12×10^−88^), and other disorders of the breast associated with childbirth and disorders of lactation (β=-3.31, p=3.18×10^−55^) are the diseases with the lowest negative iSDI associations. The top ten highest (positive effect size) and lowest (negative effect size) iSDI-disease associations are shown in Table 3. Circulatory system and digestive disease categories are the most numerous among the highest positive iSDI-disease associations, and neoplasms and sense organ disease categories are the most numerous among the lowest negative iSDI-disease associations.

**Table 3.** Top ten positive and negative iSDI-disease associations. Model specification: Disease ∼ iSDI + age + sex. Phecode disease (condition) names and categories are shown along with model coefficients.

| **Positive iSDI-disease associations** |  |  |  |  |  |
| --- | --- | --- | --- | --- | --- |
| **Disease** | **Category** | **Effect ($\beta$)** | **Std. Error** | **Z value** | **P value** |
| Schizophrenia | Mental Disorders | 5.31 | 0.08 | 63.97 | 0.00 |
| Mental retardation | Mental Disorders | 4.64 | 0.23 | 19.77 | 5.36e-87 |
| Substance addiction and disorders | Mental Disorders | 4.19 | 0.03 | 125.06 | 0.00 |
| Schizophrenia and other psychotic disorders | Mental Disorders | 3.95 | 0.07 | 54.90 | 0.00 |
| Viral hepatitis C | Infectious diseases | 3.94 | 0.06 | 64.36 | 0.00 |
| Paranoid disorders | Mental Disorders | 3.92 | 0.13 | 31.27 | 1.16e-214 |
| Poisoning by psychotropic agents | Injuries & poisonings | 3.88 | 0.10 | 38.17 | 0.00 |
| Human immunodeficiency virus [HIV] disease | Infectious diseases | 3.79 | 0.09 | 42.59 | 0.00 |
| Alcoholism | Mental Disorders | 3.60 | 0.04 | 84.17 | 0.00 |
| Hereditary disturbances in tooth structure | Digestive | 3.58 | 0.16 | 21.73 | 1.05e-104 |
| **Negative iSDI-disease associations** |  |  |  |  |  |
| **Disease** | **Category** | **Effect ($\beta$)** | **Std. Error** | **Z value** | **P value** |
| Azoospermia and oligospermia | Genitourinary | -3.46 | 0.41 | -8.34 | 7.72e-17 |
| Infertility, male | Genitourinary | -3.32 | 0.17 | -19.88 | 6.12e-88 |
| Other disorders of the breast associated with childbirth and disorders of lactation | Pregnancy complications | -3.31 | 0.21 | -15.65 | 3.18e-55 |
| Vascular hamartomas and non-neoplastic nevi | Neoplasms | -3.26 | 0.08 | -40.37 | 0.00 |
| Nevus, non-neoplastic | Neoplasms | -3.25 | 0.06 | -57.95 | 0.00 |
| Hemangioma of skin and subcutaneous tissue | Neoplasms | -3.13 | 0.06 | -54.97 | 0.00 |
| Melanomas of skin, dx or hx | Neoplasms | -3.03 | 0.14 | -21.06 | 1.81e-98 |
| Melanomas of skin | Neoplasms | -2.84 | 0.11 | -26.65 | 1.92e-156 |
| Chronic dermatitis due to solar radiation | Dermatologic | -2.78 | 0.06 | -47.89 | 0.00 |
| Actinic keratosis | Dermatologic | -2.78 | 0.06 | -47.89 | 0.00 |

We compared individual-level iSDI-disease associations to area-based zSDI-disease associations for the same 1,755 diseases (conditions), controlling for age and sex, to compare how different measures of socioeconomic deprivation affect health outcomes in the *All of Us* cohort. Overall, iSDI explains more of the variance in health outcomes than zSDI. For all diseases, iSDI incremental R2=0.008 (±0.002 95% CI) and zSDI incremental R2=0.003 (±0.0006 95% CI; Students t-test t=12.58, p=5.6×10^−35^; Figure 3A). There are 927 diseases where iSDI incremental R2 > zSDI incremental R2 compared to 684 diseases where zSDI incremental R2 > iSDI incremental R2 (Figure 3B). iSDI and zSDI show inverse patterns with respect to the number of significant positive versus negative associations with disease (Figure 3C). iSDI is positively associated with 661 diseases (37.7%), whereas zSDI is positively associated with 181 diseases (10.3%). iSDI is negatively associated with 309 diseases (17.6%), whereas zSDI is negatively associated with 664 diseases (137.8%%).

**Figure 3.**
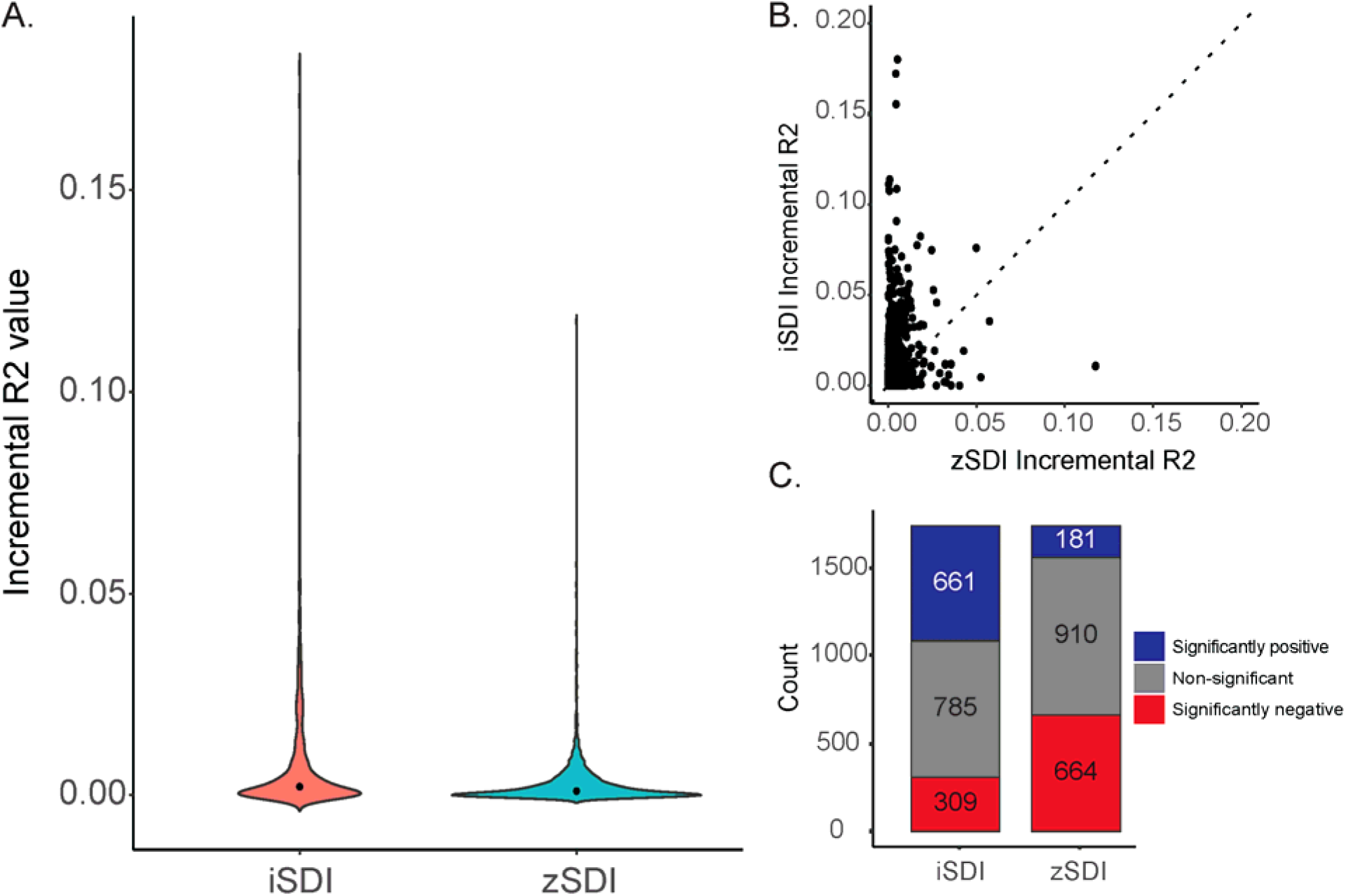
Comparison of individual-level iSDI versus area-based zSDI. (A) Distributions of incremental R2 values for iSDI (salmon) and zSDI (cyan). (B) Regression of iSDI incremental R2 values (y-axis) against zSDI incremental R2 values (x-axis). Dashed unity line is shown: above line iSDI incremental R2 > zSDI incremental R2 and below the line iSDI incremental R2 > zSDI incremental R2. (C) Numbers of significant positive (blue) and negative (red) disease associations for iSDI and zSDI.

### iSDI and health disparities

We explored a potential role for iSDI in health disparities, which are defined by the US National Institute on Minority Health and Health Disparities (NIMHD) as “health differences that adversely affect disadvantaged populations” [26]. We focused on the four largest participant self-identified race and ethnicity (SIRE) groups in the *All of Us* cohort: Asian (n=5,373, 2.65%), Black (40,262, 19.84%), Hispanic (40,762, 20.09%), and White (116,525, 57.42%; Table 1).

The Black group has the highest mean iSDI (0.55), followed by the Hispanic (0.52), White (0.29), and Asian (0.24) groups (Table 1 and Figure 4). iSDI values differ significantly across all four participant SIRE groups, with 76.8% of iSDI variation falling within groups and 23.2% of variation falling between groups (ANOVA F=20,479, p≈0; Table 4).

**Figure 4.**
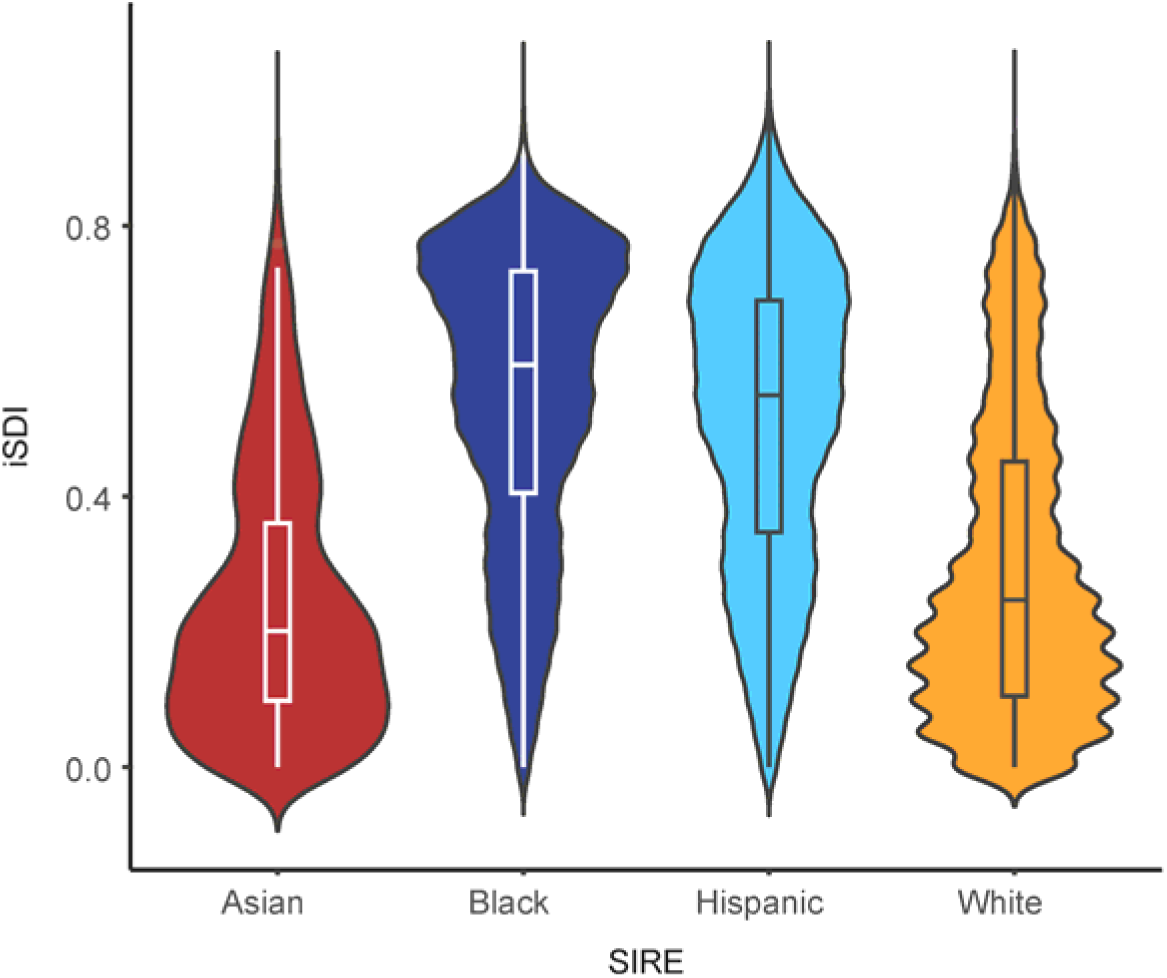
Comparison of individual-level iSDI across self-identified race and ethnicity (SIRE) groups. Distributions of iSDI are shown for the four SIRE groups studied here: Asian (red), Black (blue), Hispanic (teal), White (orange).

**Table 4.** ANOVA table for iSDI and SIRE. Groups correspond to the four SIRE groups studied here: Asian, Black, Hispanic, and White. Within (residual) and between group variation were calculated using the method of moments by equating the mean square (MS) values to the expected mean square (EMS) values. Within group (residual) EMS= V_W_ and between group EMS=n * V_W_ + V_B_, where V_W_ is the within group variation V_B_ is the between group variation, and n is the average number of individuals per group.

| **iSDI-SIRE ANOVA (F=20,479, p≈0)** |  |  |  |  |  |
| --- | --- | --- | --- | --- | --- |
|  | **df** | **SS** | **MS** | **Variation** | **%Variation** |
| **Groups** | 3 | 2,933 | 977.67 | 0.0145 | 23.24 |
| **Residuals** | 202,915 | 9,688 | 0.0477 | 0.0477 | 76.76 |

We associated SIRE with the same set of 1,755 diseases (conditions), controlling for age and sex, and identified the health disparities as diseases that are positively and significantly (Bonferroni adjusted p<2.85×10^−5^) associated with the Black and Hispanic groups, which are identified here as disadvantaged given their higher mean iSDI values. 297 diseases were found to be positively associated with the Black group and 399 were found to be positively with the Hispanic group, compared to the White reference group (Figure 5). Hypertension and diabetes were positively associated with both the Black and Hispanic groups. Hypertensive chronic kidney disease, HIV disease, substance addiction disorders, and uterine leiomyoma were positively associated with the Black group. Gingival and periodontal diseases, *H. pylori*, and pregnancy complications were positively associated with the Hispanic group.

**Figure 5.**
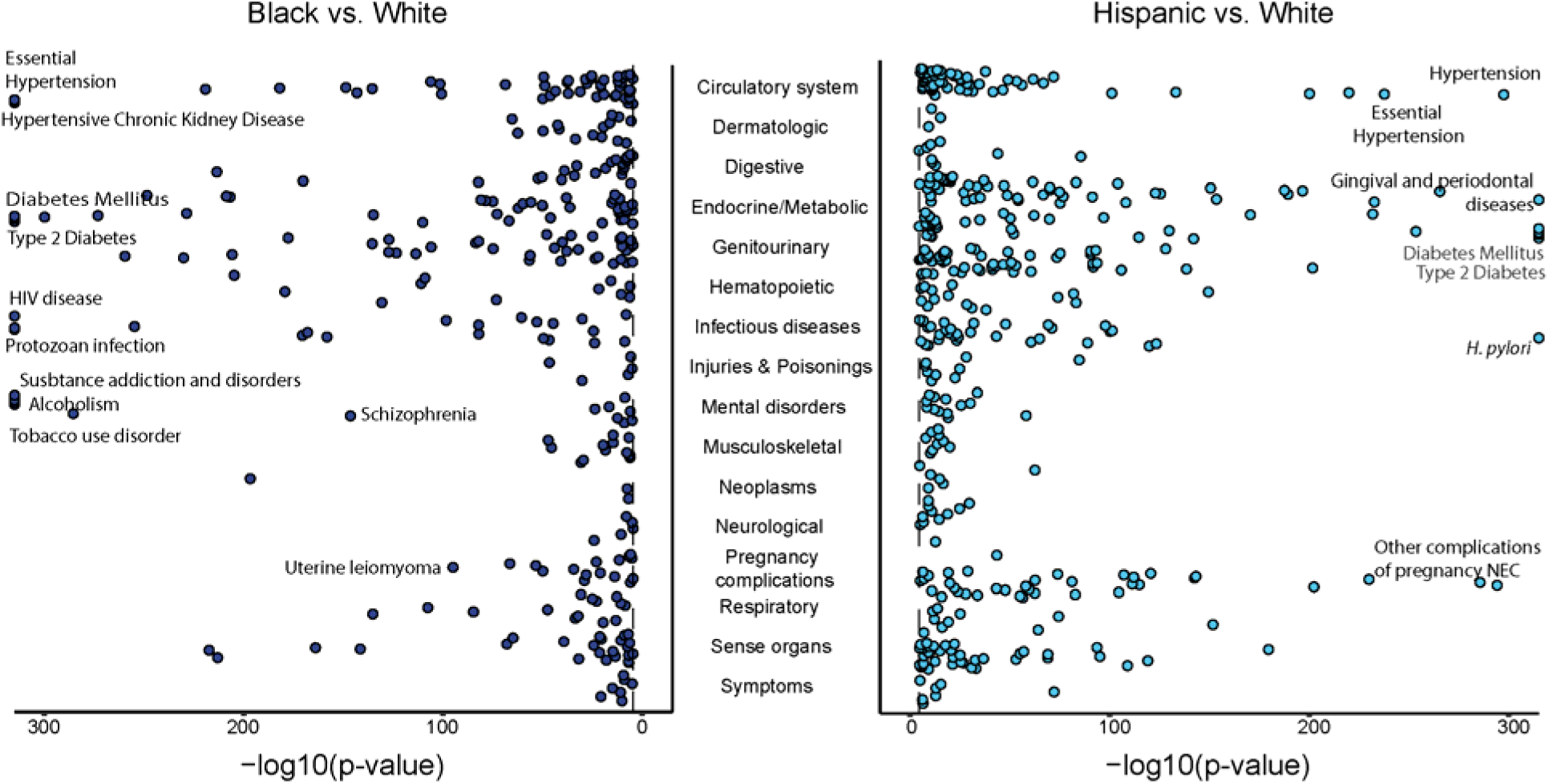
Health disparity diseases for Black (left – blue) and Hispanic (right – teal) SIRE groups. Health disparities were modeled as SIRE associations with disease (condition) outcomes, controlling for age and sex – model specification: Disease ∼ SIRE + Age + Sex – with Black and Hispanic SIRE groups compared to the White reference group. Negative log transformed p-values are shown for significantly positive disease-SIRE associations.

We evaluated the extent to which iSDI attenuates the effects of SIRE for the 399 Black and 297 Hispanic health disparity diseases. To do so, we compared the unadjusted (Disease ∼ SIRE + Age + Sex) versus iSDI adjusted (Disease ∼ SIRE + iSDI + Age + Sex) SIRE effect sizes for each health disparity disease in the Black and Hispanic groups, compared to the White reference group. For the Black group, 53.5% of diseases show significant decreases in iSDI adjusted SIRE effect sizes compared to 0.7% of diseases that show significant increases in iSDI adjusted SIRE effect sizes (Figure 6A). For the Hispanic group, 58.2% of diseases show significant decreases in iSDI adjusted SIRE effect sizes compared to 1.3% of diseases that show significant increases in iSDI adjusted SIRE effect sizes (Figure 6A).

**Figure 6.**
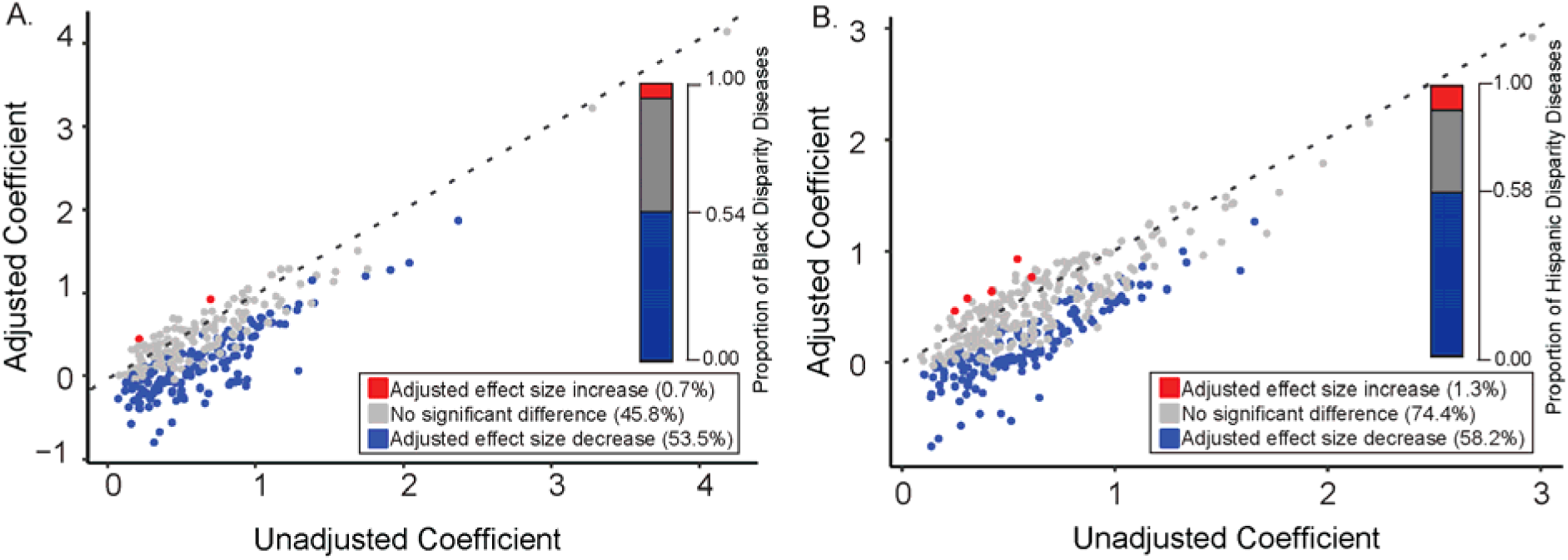
Attenuation of disease-SIRE associations by iSDI. Unadjusted SIRE effect size coefficients (x-axis) are plotted against iSDI adjusted SIRE effect size coefficients (y-axis). Unadjusted model specification: Disease ∼ SIRE + Age + Sex. iSDI adjusted model specification: Disease ∼ SIRE + iSDI + Age + Sex. Results are shown for (A) Black and (B) Hispanic health disparity diseases. Dashed unity lines are shown for each plot. Points above the unity lines indicate an increased SIRE effect size in the iSDI adjusted model, and points below the unity lines indicate a decreased SIRE effect size in the iSDI adjusted model. Numbers and proportions of health disparity diseases that show statistically significant SIRE adjusted effect size increases (red) and decreases (blue) are shown as insets in each plot.

Given the extent to which iSDI was observed to attenuate the effects of SIRE for health disparity diseases, we performed mediation analysis to evaluate whether iSDI mediates the observed disparities (Supplementary Figure 3A) [27]. Overall, the mediation analysis results are highly consistent with the results of the attenuation analysis shown in Figure 6. Specifically, the iSDI proportion attenuated values are highly correlated with the proportion mediated values from the mediation analysis (Supplementary Figure 3B & C). There are a handful of diseases with proportion attenuated and mediated values less than zero, most diseases show values between zero and one, and a substantial minority of diseases show values greater than 1. These three different ranges correspond to different kinds of mediation of SIRE effects by iSDI, related to the relative effects of SIRE versus iSDI on disease outcomes. Here, we highlight mediation analysis results for disparity diseases that show higher prevalence in Black and Hispanic participants compared to White participants, corresponding to each of the three ranges of proportion mediated values (Supplementary Table 3).

Schizophrenia shows positive total effects for both Black (0.06) and Hispanic (0.02) SIRE and proportion mediated values of 0.98 for the Black group and 1.67 for the Hispanic group (Supplementary Table 3). The indirect effect of SIRE on schizophrenia, on the mediation path through iSDI, is positive in both (0.06-Black, 0.04-Hispanic), whereas the direct effect is 0 for the Black group and –0.02 for the Hispanic group. In this case, the racial and ethnic disparities observed for schizophrenia can be attributed to higher iSDI values in those groups compared to the majority White group. For the Black group, iSDI completely mediates the observed disparity. For the Hispanic group, the indirect effect is higher than the total effect, indicating that Hispanic ethnicity has a negative association with schizophrenia when iSDI is accounted for.

Type 2 diabetes shows high positive total effects for both Black (0.14) and Hispanic (0.15) SIRE groups and proportion mediated values between zero and one (Supplementary Table 3). In both cases, the indirect effects of SIRE on disease (0.06), on the mediation path through iSDI, are only slightly lower than the direct effect (0.08-Black, 0.09-Hispanic), indicating the iSDI mediates just under half (0.43) of the observed effects of SIRE on type 2 diabetes.

Uterine leiomyoma shows positive total effects for both Black (0.04) and Hispanic (0.03) SIRE and negative proportion mediated values for each (Supplementary Table 3). The negative values for proportion mediated (−0.47-Black, –0.38 Hispanic) can be attributed to the fact that the direct effect of race and ethnicity on uterine leiomyoma is greater than the total effect, and the indirect effects are negative. Given that Black and Hispanic ethnicity are positively associated with iSDI, this means that within Black and Hispanic groups iSDI is negatively associated with uterine leiomyoma.

## DISCUSSION

Socioeconomic deprivation is recognized as an important social determinant of health, with higher levels of deprivation associated with poor health outcomes [6–11]. Here, we developed a composite metric of socioeconomic deprivation – based on education, employment, health insurance, housing, and income – for participants from the *All of Us* Research Program. Our individual-level socioeconomic deprivation index (iSDI) is complementary to the area-based metric of socioeconomic deprivation currently used by *All of Us*, which assigns deprivation values based on participants’ zip codes (zSDI) [5]. While area-based socioeconomic deprivation metrics of this kind provide valuable information about participants local environment, they may lose resolution by grouping together participants with different individual levels of deprivation. This problem is compounded by the fact that *All of Us* currently uses three-digit zip codes that may cover broad and socioeconomically heterogeneous areas.

We show that iSDI captures distinct aspects of participants’ socioeconomic deprivation and that it is significantly associated with hundreds of diseases in the *All of Us* cohort. The individual-level socioeconomic deprivation metric iSDI explains more of the variance in health outcomes for *All of Us* participants than the area-based metric zSDI, and iSDI shows substantially more positive disease associations than zSDI. Nevertheless, there are many diseases where zSDI explains more of the variance in outcomes than iSDI, indicating that these two socioeconomic deprivation metrics have complementary utility for disease modeling.

There are significant differences in the average iSDI levels among *All of Us* participant SIRE groups, with Black and Hispanic groups showing higher iSDI than White and Asian groups. Participant iSDI values are broadly distributed, however, with substantially more variation within than between SIRE groups. Health disparities are defined as differences in health outcomes that affect socially disadvantaged groups [26]. For the *All of Us* cohort, Black and Hispanic groups can be considered as socially disadvantaged, based on their higher levels of iSDI, and we found hundreds of disparity diseases associated with these groups. iSDI mediates more than half of the observed health disparities in both groups.

### Limitations

The main limitations of this study relate to the reliance on volunteer participants for the *All of Us* cohort and the use of EHR data for computational phenotyping of health outcomes. Large population biobanks, such as *All of Us* and the UK Biobank, recruit volunteer participants rather than using a population-representative sampling strategy. Volunteer participant cohorts of this kind may be biased such that they do not represent the overall population from which they are drawn, and results of the disease modeling performed here may therefore not be externally valid [14, 28, 29].

Population biobanks link participant demographic and socioeconomic data with their EHR, and we used disease diagnosis codes from EHR to create case-control cohorts for disease modeling [23]. In other words, we are modeling disease diagnosis rather than disease *per se*, which could bias results for socioeconomically deprived participants who may experience barriers to healthcare access.

### Conclusions

The composite socioeconomic deprivation index (iSDI) that we developed – based on education, employment, health insurance, housing, and income – is associated with a wide variety of health outcomes and disparities in the *All of Us* cohort. iSDI provides individual participant-level resolution on socioeconomic deprivation and is thus complementary to the area-based metric of socioeconomic deprivation currently provided for *All of Us* researchers. We make participant iSDI values available on the Researcher Workbench as a community resource in support of research on social determinants of health.

## AUTHOR CONTRIBUTIONS

Sonali Gupta and Vincent Lam: data curation, methodology, analysis, visualization, and writing. I. King Jordan and Leonardo Mariño-Ramírez: Conceptualization, funding acquisition, methodology, project administration, visualization, and writing.

## SUPPLEMENTARY MATERIAL

Supplementary Material is available at JAMIA online

## ACKNOWLEDGEMENT

We gratefully acknowledge *All of Us* participants for their contributions, without whom this research would not have been possible. We also thank the National Institutes of Health’s *All of Us* Research Program for making available the participant data analyzed in this study.

## DATA AVAILABILITY

All code and data used in this study are made available on the *All of Us* Researcher Workbench.

