## Supplemental materials for "A composite socioeconomic deprivation index from *All of Us* survey data: associations with health outcomes and disparities"

Supplementary Table 1. ***All of Us* Basics survey questions and responses used to create the individual-level socioeconomic deprivation index (iSDI).** Questions and responses are shown for education, employment, health insurance, housing, and income. The ordinal value for each question-response pair is shown, with higher orders corresponding to greater socioeconomic deprivation. The ordering shown here corresponds to the best ordering scheme out of six possible order combinations evaluated.

**Education**

***What is the highest grade or year of school you completed?***

| Order | Response |
| --- | --- |
| 8 | Never attended school or only attended kindergarten |
| 7 | Grades 1 through 4 (Primary) |
| 6 | Grades 5 through 8 (Middle school) |
| 5 | Grades 9 through 11 (Some high school) |
| 4 | Grade 12 or GED (High school graduate) |
| 3 | 1 to 3 years after high school (Some college, Associate’s degree, or technical school) |
| 2 | College 4 years or more (College graduate) |
| 1 | Advanced degree (Master’s, Doctorate, etc.) |
| na | Prefer not to answer |

**Employment**

***What is your current employment status? Please select 1 or more of these categories.***

| Order | Response |
| --- | --- |
| 4 | Unable to work (disabled) |
| 4 | Out of work for 1 year or more |
| 3 | Out of work for less than 1 year |
| 3 | A student |
| 2 | Self-employed |
| 1 | Retired |
| 1 | Employed for wages (part- time or full-time) |
| na | Skip |
| na | Prefer not to answer |
| na | A homemaker |

**Housing**
***Do you own or rent the place where you live in?***

| Order | Response |
| --- | --- |
| 3 | Skip |
| 2 | Rent |
| 1 | Own |
| na | Prefer not to answer |
| na | Don’t know |
| na | Other arrangement |

**Income**
***What is your annual household income from all sources?***

| Order | Response |
| --- | --- |
| 9 | Less than $10,000 |
| 8 | $10,000- $24,999 |
| 7 | $25,000-$34,999 |
| 6 | $35,000-$49,999 |
| 5 | $50,000- $74,999 |
| 4 | $75,000-$99,999 |
| 3 | $100,000- $149,999 |
| 2 | $150,000- $199,999 |
| 1 | $200,000 or more |
| na | Prefer not to answer |
| na | Skip |

**Insurance**
***Are you covered by health insurance or some other kind of health care plan?***

| Order | Response |
| --- | --- |
| 2 | No |
| 2 | Prefer not to answer |
| 2 | Don’t know |
| 1 | Yes |
| na | Skip |

Supplementary Table 2. **Survey question-response ordering schemes.** Responses to *All of Us* participant survey questions on education, employment, health insurance, housing, and income were ordered with higher values corresponding to greater socioeconomic deprivation (Supplementary Table 1). Since there was some ambiguity regarding how responses could be ordered from lowest to highest socioeconomic deprivation for questions on employment and insurance, six different ordering schemes were evaluated, as shown here. Answers that show ‘na’ were not used.

| **Employment** | **Order 1** | **Order 2** | **Order 3** | **Order 4** | **Order 5** | **Order 6** |
| --- | --- | --- | --- | --- | --- | --- |
| Employment Status: Unable To Work | 4 | 4 | 4 | 4 | 4 | 4 |
| Employment Status: Out Of Work One Or More | 4 | 4 | 4 | 4 | 4 | 4 |
| Employment Status: Student | 3 | 4 | 3 | 4 | 3 | 4 |
| Employment Status: Out Of Work Less Than One | 3 | 3 | 3 | 3 | 3 | 3 |
| Employment Status: Self Employed | 2 | 2 | 2 | 2 | 2 | 2 |
| Employment Status: Retired | 1 | 2 | 1 | 2 | 3 | 1 |
| Employment Status: Employed For Wages | 1 | 1 | 1 | 1 | 1 | 1 |
| PMI: Skip | na | na | na | na | na | na |
| PMI: Prefer Not To Answer | na | na | na | na | na | na |
| Employment Status: Homemaker | na | na | na | na | na | na |
| **Insurance** |  |  |  |  |  |  |
| Health Insurance: No | 2 | 2 | 2 | 2 | 2 | 2 |
| PMI: Prefer Not to answer | 2 | 2 | 2 | 2 | 2 | 2 |
| PMI: Dont know | 2 | 2 | NA | NA | 2 | 2 |
| Health Insurance: Yes | 1 | 1 | 1 | 1 | 1 | 1 |
| PMI: Skip | na | na | na | na | na | na |

Supplementary Figure 1. **Comparison of question-response ordering schemes.** Responses to *All of Us* participant survey questions on education, employment, health insurance, housing, and income were ordered with higher values corresponding to greater socioeconomic deprivation (Supplementary Table 1). Since there was some ambiguity regarding how responses could be ordered from lowest to highest socioeconomic deprivation for different questions, six different ordering schemes were evaluated. Spearman correlation values for all pairs of questions-responses were calculated for each ordering. The median Spearman correlation values for the six different orderings are shown. The ‘Best order’ corresponds to Order 1 in Supplementary Table 2.

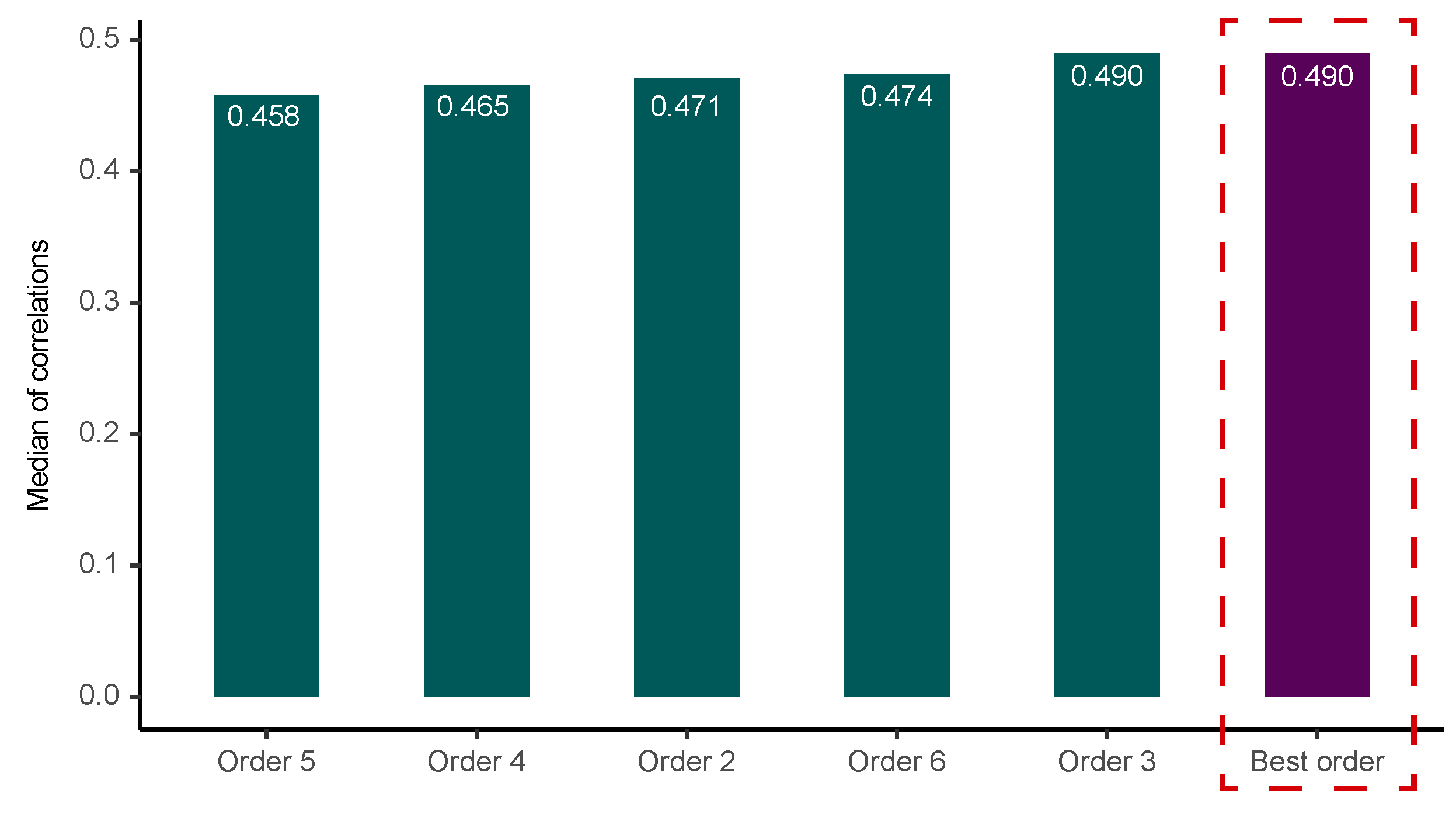

Supplementary Figure 2. **Pairwise Spearman rank (ρ) correlations between the five constituent socioeconomic deprivation measures taken from the *All of Us* participant survey.** Values shown here correspond to correlations calculated prior to imputation of missing response data, compared to the values shown in Figure 1A, which correspond to correlations calculated after imputation.

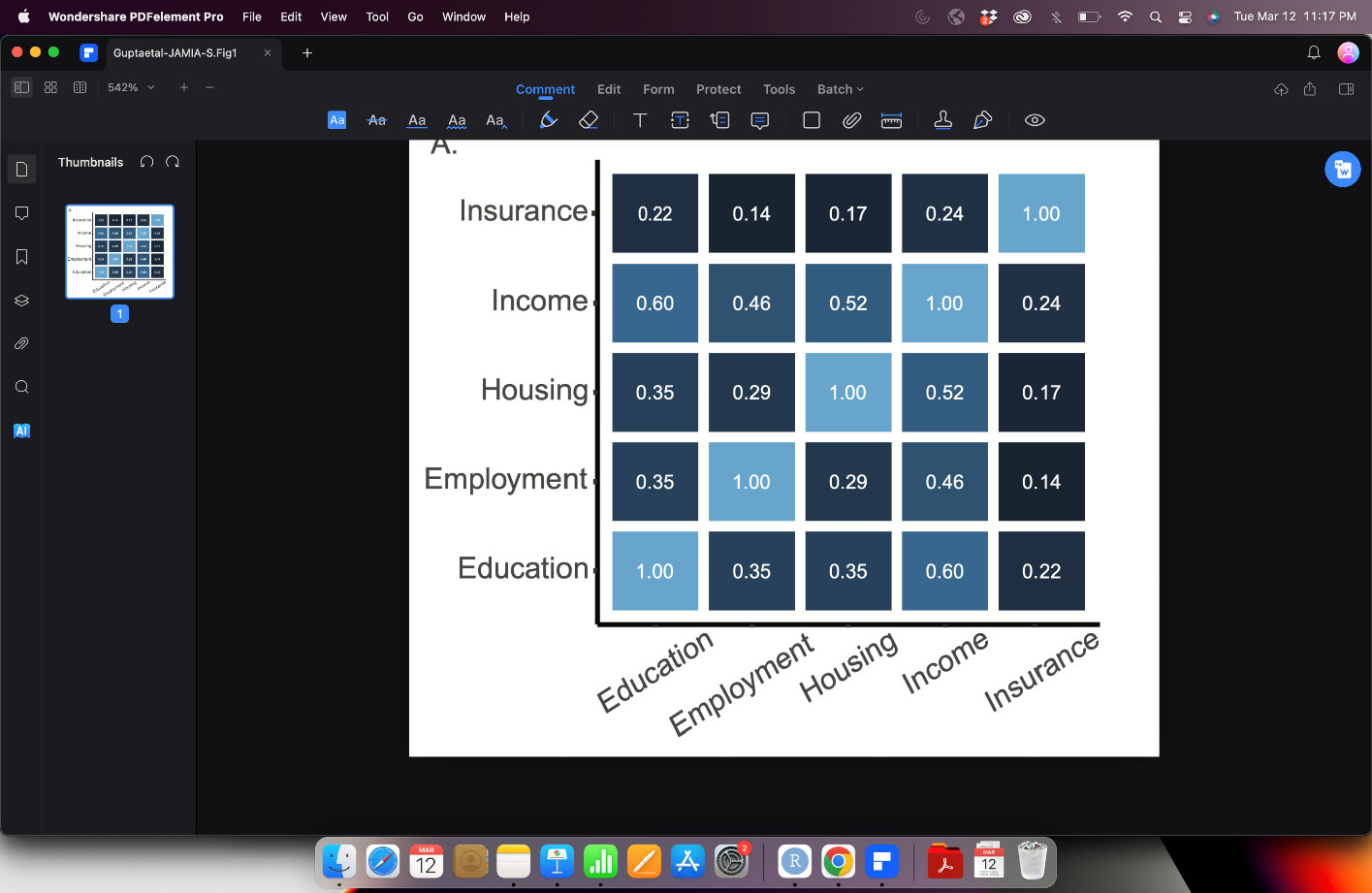

Supplementary Figure 3. **Comparison between iSDI mediation and percent attenuation analyses.** (A) Directed acyclic graph showing the iSDI mediation (indirect effects) of participant self-identified race and ethnicity (SIRE) associations (direct effect) with disease outcomes. (B and C). Proportion of the SIRE effect size that is mediated (y-axis) compared to the attenuation proportion (x-axis) shown for Black and Hispanic disparity diseases.

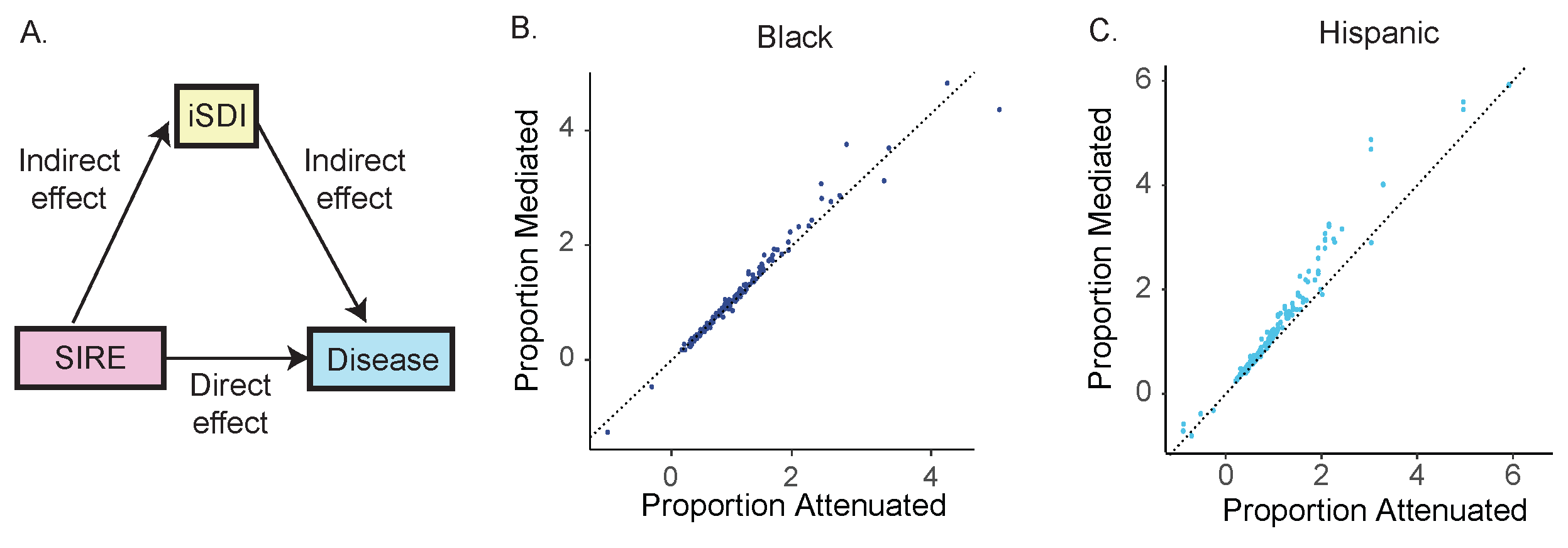

Supplementary Table 3. **Mediation analysis for iSDI and health disparities.** SIRE is the exposure, iSDI is the mediator, and disease is the outcome. The total effect is the sum of the indirect and direct effects of SIRE on disease, where the indirect effect is mediated by iSDI (Figure 7A). The proportion mediated (Prop. Med.) measures the proportion of the total effect of SIRE on disease mediated by iSDI.

|  | **Black** | | | **Hispanic** | | |
| --- | --- | --- | --- | --- | --- | --- |
|  | **Estimate** | **95% CI upper, lower** | **P value** | **Estimate** | **95% CI upper, lower** | **P value** |
|  | **Schizophrenia** | | | | | |
| **Total effect** | 0.06 | 0.06, 0.06 | <2e-16 | 0.02 | 0.02, 0.02 | <2e-16 |
| **Indirect effect** | 0.06 | 0.06, 0.06 | <2e-16 | 0.04 | 0.03, 0.04 | <2e-16 |
| **Direct effect** | 0.00 | 0.00, 0.00 | 0.46 | -0.02 | -0.02, -0.01 | <2e-16 |
| **Prop. Med.** | 0.98 | 0.92, 1.03 | <2e-16 | 1.90 | 1.67, 2.19 | <2e-16 |
|  | **Type 2 diabetes** | | | | | |
| **Total effect** | 0.14 | 0.13, 0.14 | <2e-16 | 0.15 | 0.14, 0.16 | <2e-16 |
| **Indirect effect** | 0.06 | 0.06, 0.06 | <2e-16 | 0.06 | 0.06, 0.07 | <2e-16 |
| **Direct effect** | 0.08 | 0.07, 0.08 | <2e-16 | 0.09 | 0.08, 0.09 | <2e-16 |
| **Prop. Med.** | 0.43 | 0.42, 0.46 | <2e-16 | 0.43 | 0.41, 0.45 | <2e-16 |
|  | **Uterine leiomyoma** | | | | | |
| **Total Effect** | 0.04 | 0.04, 0.04 | <2e-16 | 0.03 | 0.02, 0.03 | <2e-16 |
| **Indirect effect** | -0.04 | -0.02, -0.02 | <2e-16 | -0.01 | -0.01, -0.01 | <2e-16 |
| **Direct effect** | 0.06 | 0.06, 0.06 | <2e-16 | 0.04 | 0.03, 0.04 | <2e-16 |
| **Prop. Med.** | -0.47 | -0.52, -0.42 | <2e-16 | -0.38 | -0.46, -0.32 | <2e-16 |
